# Percutaneous assessment of the relationship between Homologous and Non-homologous bones of Upper and Lower limbs among Ghanaian tertiary students

**DOI:** 10.1101/2024.12.14.24319029

**Authors:** Daniel Kobina Okwan, Chrissie Stansie Abaidoo, Juliet Robertson, Samuel Kwadwo Peprah Bempah, Pet-Paul Wepeba, Thomas Kwaku Asante, Priscilla Obeng, Ethel Akua Achiaa Domfeh, Sarah Owusu Afriyie

**Author notes:** Corresponding author (DKO).

## Abstract

**Background:** Natural disasters and accidents may result in loss of some part of the upper and or lower limbs. In such situations, there is the need to design appropriate best fit prostheses for such victims. There is the need to have sex-and side-specific models that can be applied conveniently to design appropriate prosthesis for maximum benefit to the patient.

**Aim:** Therefore, this study was designed to determine the relationship between homologous and non-homologous bones of upper and lower limbs among Ghanaians using a percutaneous approach.

**Methodology:** Ethical approval for this cross-sectional study was sought from the Committee on Human Research, Publication and Ethics, KNUST and 242 participants were sampled via a purposive sampling technique after seeking their consent.

**Results:** Males significantly recorded greater percutaneous long bones for both the upper and lower limbs than the female participants. With the exception of radial length among the females, for both sexes bilateral asymmetry was observed in the remaining percutaneous limb dimensions where the upper limb parameters were greater on the left side than the right side, but showed right dominance for the lower limb parameters. Although several useful equations were derived, most of them utilized humeral length in the prediction of lower limb length estimation.

**Conclusion:** Findings of the present study are useful for the biological profiling of humans with dismembered body parts involved in various disasters such as automobile accidents. The formulae derived would be useful for the design of appropriate prostheses. This study has also reaffirmed the existence of sexual and bilateral dimorphism in body dimensions.

## Introduction

Scientists including anatomists, anthropologists and forensic medicine experts have immense interest in anthropometric studies [1], including long bones. Endogenous variables including genetic, hormonal, metabolic and target tissue responses are some of the elements influencing bone formation and therefore its dimensions and configuration. Numerous local, regional and physical factors can affect bone length [2]. According to Dumitru *et al.* [3], external variables that can also alter bone formation include diet, exercise and psychosocial factors. Long bone formation has been linked to sex hormones [4].

Due to the difference in the oestrogen-to-testosterone ratio, particularly at the commencement of puberty, a person’s sex also affects their limb proportions [5,6]. The nature of occupation or task one is involved in, lifestyle particularly due to the influence of civilisation may be impactful on body parts, including the limbs [7]. Carlson and Marchi [8] also asserted that, in addition to the environment, physical stress, body composition and hormones also have an impact on bone structure, particularly that of the lower limb. Moreover, it has been reported that, reduced leg length with respect to the torso may be implicated by heavy mechanical stress, trauma, infectious diseases and perhaps intrauterine poor nutrition to the developing foetus [9]. Also, data obtained from body measurements with respect to race, age and sex could be utilized in the design of products including prostheses [10,11]. The challenge for medicolegal specialists is identification using dismembered body parts of the deceased [12]. These dismembered body parts arise as a result of natural disasters (including earthquakes, tsunamis and floods) and man-induced disasters (bomb attack by terrorists, vehicular accidents and wars) or even attack by wild animals [13]. Additionally, in such predicaments, there could be damage to biological identifiable areas such as the face, fingers and eyes [14,15].

In fact, the events of natural calamities continue to increase every now and then with mutilated body parts frequently encountered [16]. Specifically, in Ghana, reports show that there is increased natural disasters and road traffic accidents resulting in multiple casualties [17].

Therefore, there is the utmost need to obtain standard formulae for the assessment of the relationship between the homologous and non-homologous bones of the upper and lower limbs. Moreover, the obtained relationship between the long bones would be useful for the design of best-fit prostheses for amputated upper and or lower limbs. Therefore, in order to develop regression formulae for the link between the homologous and non-homologous percutaneous bones of the upper and lower limbs for diverse applications, the present study sought to analyze and throw more light on the association between percutaneous lengths of the limbs among Ghanaian tertiary using a percutaneous approach. Specifically, the study sought to:

- measure percutaneous long bone lengths of the study participants
- evaluate the level of sexual and bilateral dimorphism among the measured long bones
- assess the relationship between homologous and non-homologous bones of upper and lower limbs and determine the best predictors among the upper limb bones for lower limb bone length estimation

## Materials and Methods

### Study Design and Ethical Consideration

This study employed a cross-sectional approach to recruit participants aged 20 – 25 years. Ethical approval was sought from Committee on Human Research, Publication and Ethics, School of Medical Sciences, Kwame Nkrumah University of Science and Technology (KNUST), Kumasi, Ghana. All procedures performed on the participants were done in accordance with the ethical standards outlined in the 1964 and subsequent modifications of the Declaration of Helsinki. Informed consent was as well sought from each study participant before relevant data were taken confidentially. A purposive sampling technique was employed to recruit a total of 242 participants.

### Inclusion and Exclusion Criteria

Only consented Ghanaian tertiary students aged 20 - 25 years were recruited for the present study. Individuals with old or current fractures of the upper or lower limb or other orthopaedic deformities that could affect the measurements of the long bones were excluded. Also, individuals with metabolic or developmental disorders than could potentially affect the maximum dimensions of the bones were excluded from the present study.

### Measurements Taken

**Humeral length:** Percutaneous distance from the lateral end of the clavicle to the lateral epicondyle of the humerus [18]. This was done with the elbow bent at an angle of 90° and participants being in siting position and the shoulder fully adducted.

**Ulnar length:** It was measured as the distance from the tip of the olecranon process of the ulna to the head of the ulna [19]. In other words, it is the displacement of the styloid process from the tip of the olecranon process with the elbow fully flexed (90°) and palm spread over the opposite shoulder.

**Radial length**: It was measured as the displacement from the radial head to the radial styloid process.

**Femoral length**: It was measured as the displacement from the greater trochanter to the lateral epicondyle of the femur.

**Tibial length:** It was measured as the distance from the medial-most superficial point on the upper border of the medial condyle of the tibia to the tip of the medial malleolus (Spherion) of the tibia [20]. The angle between flexor surface of the leg and the thigh was maintained at 90°. This was achieved by asking each participant to sit (as it is easier to access the tibia in this position) facing the observer with ankle resting on the knee, so that the medial aspect of the tibia faced upwards.

**Fibular length:** It was measured as the distance from the fibular head to the lateral malleolus. Each participant was asked to sit in order to make the landmarks easily accessible and palpable.

### Precautions observed during measurements

To reduce both inter-observer and intra-observer rate of error and to ensure reproducibility, all measurements were taken twice by the same investigator. Long bone lengths of all participants were taken at a specific time from 10 am to 2 pm. Some of the landmarks were highlighted on the skin using a marking pencil to ensure accuracy.

### Definition of Key Concepts

**Long bones** refer to those that are longer than they are wide and are found in the upper and lower limbs of the human body.

**Homologous bones** refer to those that arise from the same embryonic origin although perform different functions and are found in different parts of the limbs (upper and lower). Therefore, the homologous bones of the upper and lower limbs are humerus and femur; radius and tibia; ulna and fibula.

**Non-homologous bones** refer to those that arise from different embryonic origins and thus do not share a common embryonic origin. Thus, for the upper and lower limbs, humerus is non-homologous with tibia and fibula. For the radius, it is non-homologous with the femur and fibula. Also, the ulna is non-homologous with the femur and tibia.

**Relationship** refers to the correspondence or connection between two or more variables.

**Prosthesis** refers to an artificial body part such as a limb.

### Statistical Analysis

Data obtained were entered into Microsoft Office Excel Spreadsheet 2019 and analyzed using IBM SPSS version 26.0 software. For descriptive statistics, data were expressed in means alongside standard deviation. Independent samples t-test was employed to assess sexual dimorphism in the study participants based on parameters studied. Existence of bilateral asymmetry was assessed using paired samples t-test. Statistically significant level for differences or otherwise was pegged at p < 0.05 (95% confidence level). Pearson’s correlation coefficient (r) was used as the measure of strength and direction of the association between percutaneous homologous and non-homologous bone lengths for both male and female participants. Coefficient of determination (R^2^) together with its adjusted value (adjusted R^2^) was estimated to determine how much of the variance in the dependent variable could be explained by its relationship to the other variables. Using stepwise approach, linear regression equations were derived as predictive models for the prediction of lower limb bone lengths using upper limb bone measurements.

## Results

### Descriptive statistics

The mean age of the male participants was 21.58 ± 1.70 years with that of the females being 21.49 ± 1.64 years. As shown in Tables 1 and 2, although sexual dimorphism was observed in the measured parameters (p <0.001), for both sexes, regarding the anthropometric parameters of upper limb bone length measurements, there was a gradual decrease from humeral length, through ulnar length and then to radial length. For the lower limb bones however, the length of the bones declined gradually from femoral length, through tibial length and then to fibular length.

**Table 1:**
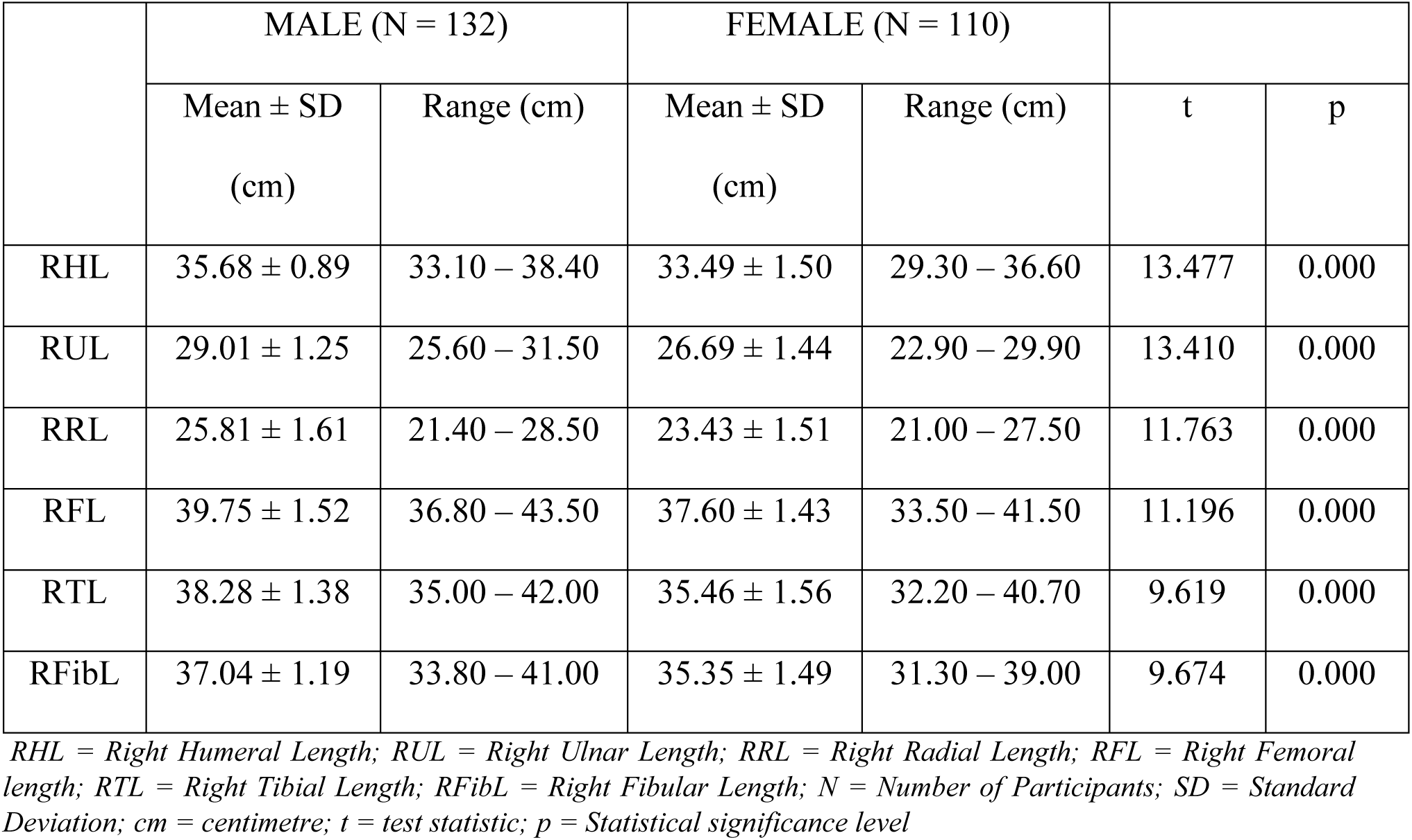
Descriptive Statistics of Right Long Bone Lengths of Study Participants Stratified by Sex.

**Table 2:** Descriptive Statistics of Left Long Bone Lengths of Study Participants Stratified by Sex.

|  | MALE (N = 132) |  | FEMALE (N = 110) |  |  |  |
| --- | --- | --- | --- | --- | --- | --- |
| | Mean ± SD<br>(cm) | Range (cm) | Mean ± SD<br>(cm) | Range (cm) | t | p |
| LHL | 35.85 ± 0.93 | 33.50 – 39.00 | 33.60 ± 1.57 | 29.50 – 37.80 | 13.185 | 0.000 |
| LUL | 29.10 ± 1.27 | 25.80 – 31.70 | 26.78 ± 1.51 | 23.20 – 30.80 | 12.997 | 0.000 |
| LRL | 25.96 ± 1.60 | 21.70 – 27.80 | 23.46 ± 1.58 | 20.80 – 27.10 | 12.154 | 0.000 |
| LFL | 39.45 ± 1.56 | 36.10 – 43.30 | 37.37 ± 1.46 | 32.50 – 41.20 | 10.784 | 0.000 |
| LTL | 38.00 ± 1.50 | 34.60 – 42.80 | 36.29 ± 1.52 | 32.00 – 40.80 | 8.963 | 0.000 |
| LFibL | 36.86 ± 1.28 | 33.40 – 42.00 | 35.16 ± 1.53 | 31.00 – 38.80 | 9.281 | 0.000 |

*LHL = Left Humeral Length; LUL = Left Ulnar Length; LRL = Left Radial Length; LFL = Left Femoral length; LTL = Left Tibial Length; LFibL = Left Fibular Length; N = Number of Participants; SD = Standard Deviation; cm = centimetre; t = test statistic; p = Statistical significance level*

### Assessment of Bilateral Asymmetry of Study Participants

Bilateral asymmetry in males was seen in all the measured parameters (p < 0.05). For the females, with the exception of radial length, bilateral asymmetry was observed in the remaining parameters (p < 0.05) (Table 3).

**Table 3:**
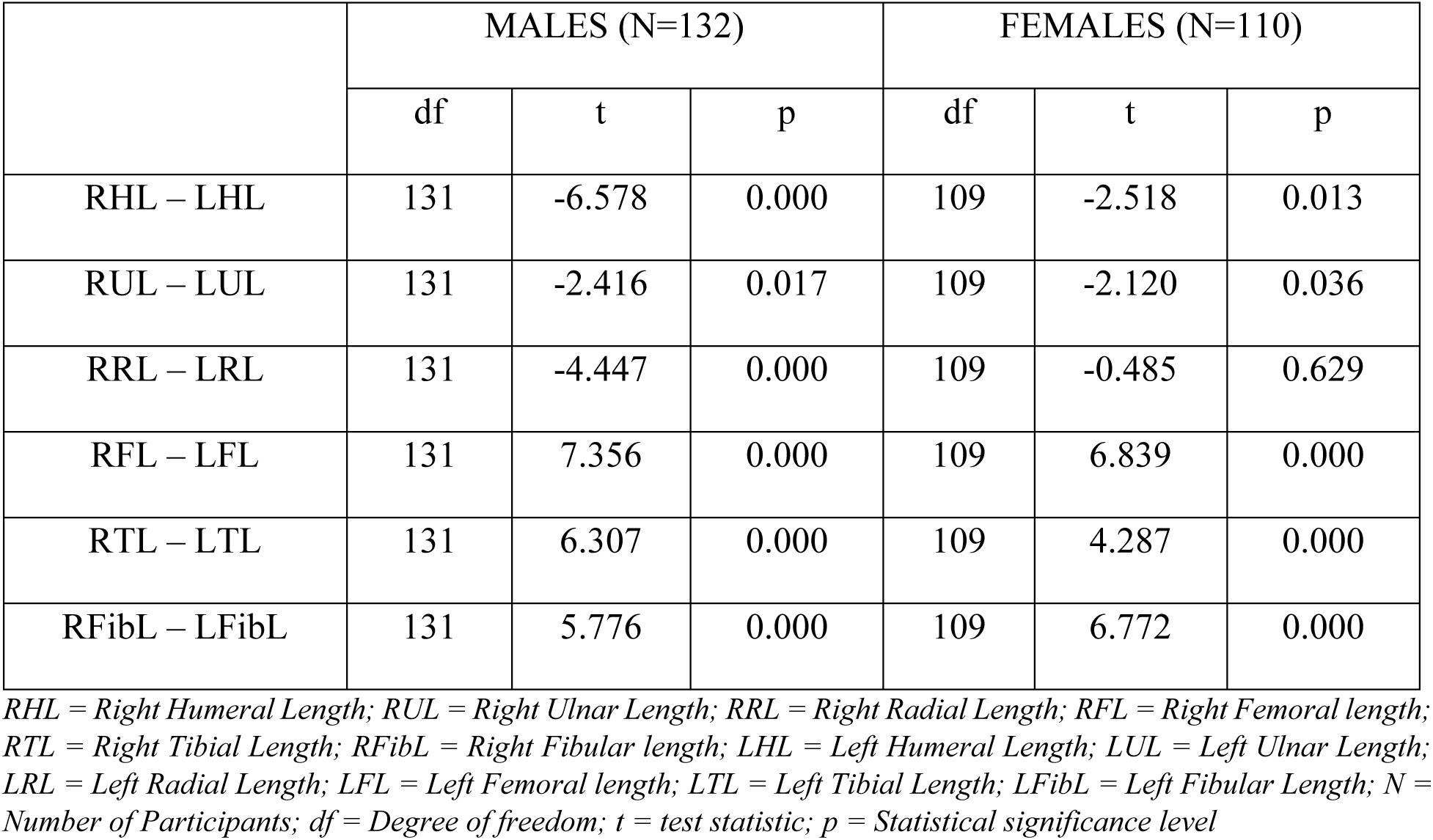
Comparison of Right and Left Anthropometric Parameters of Study Participants Stratified by Sex.

|  | MALES (N=132) |  |  | FEMALES (N=110) |  |  |
| --- | --- | --- | --- | --- | --- | --- |
|  | df | t | p | df | t | p |
| RHL – LHL | 131 | -6.578 | 0.000 | 109 | -2.518 | 0.013 |
| RUL – LUL | 131 | -2.416 | 0.017 | 109 | -2.120 | 0.036 |
| RRL – LRL | 131 | -4.447 | 0.000 | 109 | -0.485 | 0.629 |
| RFL – LFL | 131 | 7.356 | 0.000 | 109 | 6.839 | 0.000 |
| RTL – LTL | 131 | 6.307 | 0.000 | 109 | 4.287 | 0.000 |
| RFibL – LFibL | 131 | 5.776 | 0.000 | 109 | 6.772 | 0.000 |

*RHL = Right Humeral Length; RUL = Right Ulnar Length; RRL = Right Radial Length; RFL = Right Femoral length; RTL = Right Tibial Length; RFibL = Right Fibular length; LHL = Left Humeral Length; LUL = Left Ulnar Length; LRL = Left Radial Length; LFL = Left Femoral length; LTL = Left Tibial Length; LFibL = Left Fibular Length; N = Number of Participants; df = Degree of freedom; t = test statistic; p = Statistical significance level*

### Correlation Between Percutaneous Bone Lengths of Upper and Lower Limbs of Study Participants Stratified by Sex and Laterality

Table 4 shows the Pearson’s correlation between homologous and non-homologous bone lengths of upper and lower limbs taking laterality and sex into consideration.

**Table 4:**
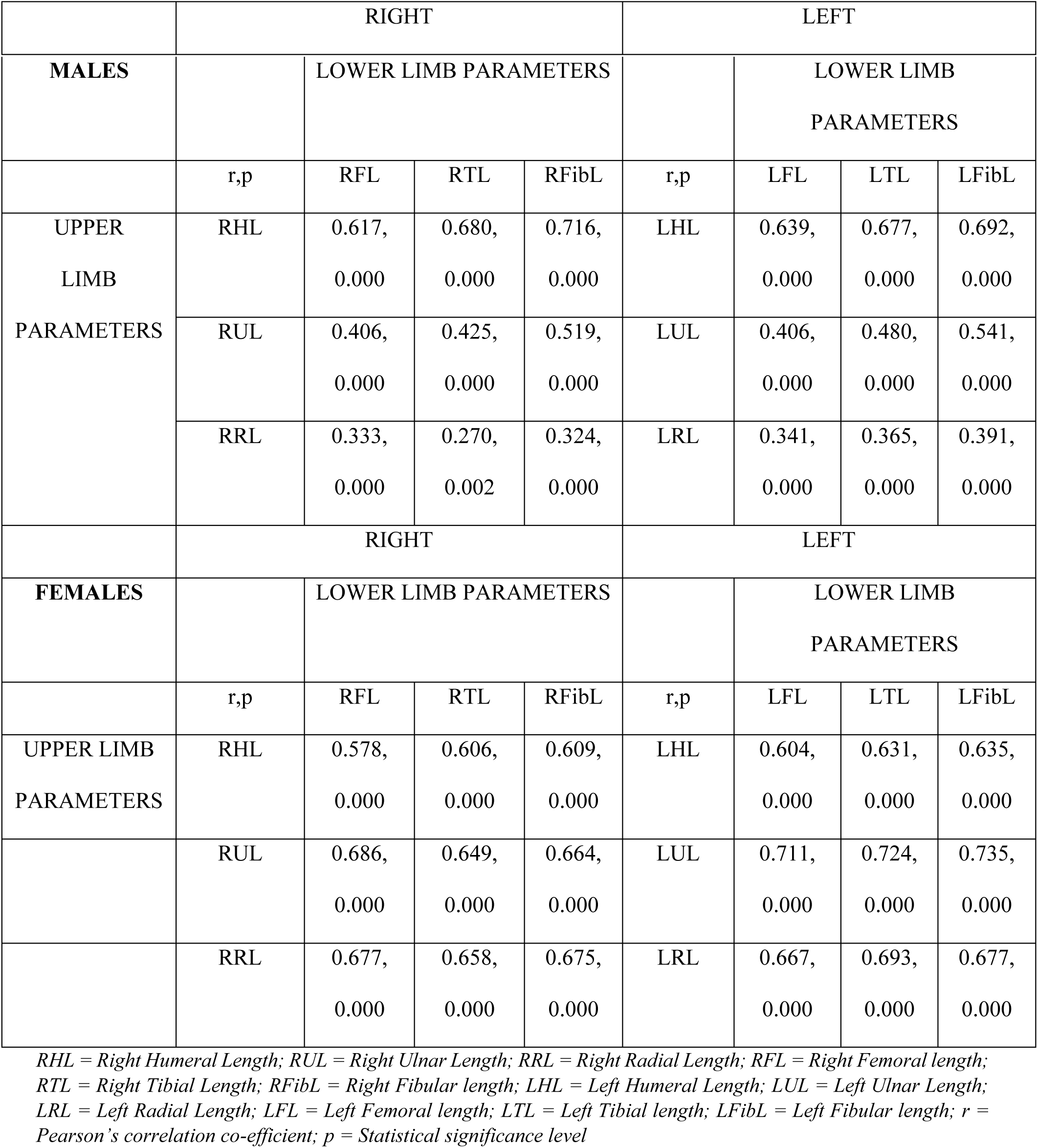
Pearson’s Correlation Between Upper and Lower Limb Bone Lengths of Study Participants Stratified by Sex and Laterality.

### Regression Models for Lower Limb Bone Lengths Estimation Using Lengths of Upper Limb Bones of the Study Participants based on Sex and Laterality

Linear regression analysis for estimation of lower limb percutaneous bone lengths using that of upper limb was done for homologous and non-homologous bone lengths considering the male (Table 5) and female (Table 6) study participants.

**Table 5:** Linear Equations for Estimation of Lower Limb Bone Lengths Using That of Upper Limb for The Male Study Participants.

| MALE PARTICIPANTS |  |  |  |
| --- | --- | --- | --- |
| EQUATION | R<sup>2</sup> | ADJ.R<sup>2</sup> | SEE |
| **RIGHT** |  |  |  |
| **RFL = 2.040 + 1.056 (RHL)** | 0.381 | 0.376 | 1.2012 |
| RFL = 0.884 + 0.973 (RHL) + 0.160 (RRL) | 0.407 | 0.398 | 1.1798 |
| **RTL = 32.302 + 0.231 (RRL)** | 0.073 | 0.066 | 1.3381 |
| RTL = 0.430 + 1.061 (RHL) | 0.463 | 0.459 | 1.0184 |
| **RFibL = 22.756 + 0.492 (RUL)** | 0.270 | 0.264 | 1.0205 |
| RFibL = 2.823 + 0.959 (RHL) | 0.513 | 0.509 | 0.8334 |
| RFibL = 1.777 + 0.812 (RHL) + 0.216 (RUL) | 0.553 | 0.546 | 0.8015 |
| **LEFT** |  |  |  |
| **LFL = 1.100 + 1.071 (LHL)** | 0.409 | 0.404 | 1.2073 |
| **LTL = 29.430 + 0.330 (LRL)** | 0.133 | 0.127 | 1.3544 |
| LTL = 0.349 + 1.050 (LHL) | 0.458 | 0.454 | 1.0707 |
| LTL = -0.365 + 0.909 (LHL) + 0.198 (LUL) | 0.480 | 0.472 | 1.0527 |
| **LFibL = 21.023 + 0.544 (LUL)** | 0.293 | 0.288 | 1.0810 |
| LFibL = 2.844 + 0.949 (LHL) | 0.479 | 0.475 | 0.9280 |
| LFibL = 1.946 + 0.772 (LHL) + 0.249 (LUL) | 0.524 | 0.516 | 0.8908 |

*RHL = Right Humeral Length; RUL = Right Ulnar Length; RRL = Right Radial Length; RFL = Right Femoral Length; RTL = Right Tibial Length; RFibL = Right Fibular Length; LHL = Left Humeral Length; LUL = Left Ulnar Length; LRL = Left Radial Length; LFibL = Left Fibular length; LTL = Left Tibial Length; ADJ. = Adjusted; R<sup>2</sup> = Regression co-efficient of determination; SEE = Standard Error of Estimate; r = Pearson's correlation co-efficient; p = Statistical significance level; Equations involving solely homologous bones are indicated in bold*

**Table 6:** Linear Equations for Estimation of Percutaneous Lower Limb Bone Lengths Using That of Upper Limb for The Female Study Participants.

| FEMALE PARTICIPANTS |  |  |  |
| --- | --- | --- | --- |
| EQUATION | R<sup>2</sup> | ADJ.R<sup>2</sup> | SEE |
| **RIGHT** |  |  |  |
| **RFL = 19.187 + 0.550 (RHL)** | 0.334 | 0.328 | 1.1699 |
| RFL = 19.451 + 0.680 (RUL) | 0.471 | 0.466 | 1.0428 |
| RFL = 18.184 + 0.410 (RUL) + 0.362 (RRL) | 0.542 | 0.534 | 0.9745 |
| RFL = 13.302 + 0.233 (RUL) + 0.377 (RRL) + 0.276 (RHL) | 0.599 | 0.587 | 0.9170 |
| **RTL = 20.470 + 0.682 (RRL)** | 0.433 | 0.428 | 1.1794 |
| RTL = 9.932 + 0.518 (RRL) + 0.430 (RHL) | 0.579 | 0.571 | 1.0210 |
| **RFibL = 17.072 + 0.685 (RUL)** | 0.440 | 0.435 | 1.1168 |
| RFibL = 19.732 + 0.666 (RRL) | 0.456 | 0.451 | 1.1016 |
| RFibL = 9.775 + 0.511 (RRL) + 0.406 (RHL) | 0.599 | 0.592 | 0.9498 |
| **LEFT** |  |  |  |
| **LFL = 18.489 + 0.562 (LHL)** | 0.365 | 0.359 | 1.1706 |
| LFL = 18.896 + 0.690 (LUL) | 0.505 | 0.500 | 1.0333 |
| LFL = 18.078 + 0.466 (LUL) + 0.291 (LRL) | 0.551 | 0.543 | 0.9887 |
| LFL = 13.583 + 0.253 (LUL) + 0.330 (LRL) + 0.276 (LHL) | 0.603 | 0.591 | 0.9344 |
| **LTL = 20.736 + 0.663 (LRL)** | 0.480 | 0.475 | 1.0982 |
| LTL = 16.779 + 0.728 (LUL) | 0.524 | 0.519 | 1.0507 |
| LTL = 15.836 + 0.470 (LUL) + 0.335 (LRL) | 0.581 | 0.573 | 0.9906 |
| LTL = 10.611 + 0.223 (LUL) + 0.381 (LRL) + 0.320 (LHL) | 0.646 | 0.636 | 0.9149 |
| **LFibL = 15.219 + 0.745 (LUL)** | 0.540 | 0.536 | 1.0386 |
| LFibL = 10.865 + 0.564 (LUL) + 0.274 (LHL) | 0.588 | 0.580 | 0.9879 |
| LFibL = 9.344 + 0.283 (LUL) + 0.311 (LHL) + 0.332 (LRL) | 0.642 | 0.632 | 0.9252 |

*RHL = Right Humeral Length; RUL = Right Ulnar Length; RRL = Right Radial Length; RFL = Right Femoral Length; RTL = Right Tibial Length; RFibL = Right Fibular Length; LHL = Left Humeral Length; LUL = Left Ulnar Length; LRL = Left Radial Length; LFibL = Left Fibular length; LTL = Left Tibial Length ADJ. = Adjusted; R<sup>2</sup> = Regression co-efficient of determination; SEE = Standard Error of Estimate; r = Pearson's correlation co-efficient; p = Statistical significance level; Equations involving solely homologous bones are indicated in bold*

## Discussion

### Descriptive statistics of percutaneous bones of upper and lower limbs

There was a progressive decrease in length starting with the femoral length and continuing through the tibial length, fibular length, humeral length, ulnar length and finally the radial length for both the right and left percutaneous long bones of the upper and lower limbs (Tables 1 and 2). This trend is expected as the lower limb is evidently more extended than the upper limb and that regarding the lower limb, the longest bone is the femur, with that of the upper limb being the humerus. In addition, the ulna is longer than the radius with the weight-bearing tibia being longer than the non-weight bearing fibula. Since this is the usual anatomical trend in proportions of the limb lengths, it suggests normal distribution pattern in long bones regardless of sex. This finding holds agreement with the fact that, despite the variations among populations, there are important consistent relationships between body parts and the entire body as a whole [21]. Percutaneous evaluation of long bone lengths has been done by other investigators on different populations [16,22–28]. Malli *et al.* [22] employed only arm length for stature estimation, whereas Mehta *et al.* [24] utilized only tibial length for stature estimation among a population of Central India. In a Nigerian population, Nandi *et al.* [25] worked on bones of the upper limb for stature reconstruction. Panjakash *et al.* [26] utilized the length of the forearm for stature estimation among the people of North Karnataka While Sukumar [16] employed only ulnar length, Sume [28] utilised only tibial length for stature prediction among their respective study populations. For the people of Jordan, Mashali *et al.* [23] employed measurements of the extremities for stature estimation. Unlike the aforementioned studies, the present study used lengths of the upper limb to predict that of the lower limb for various forensic and medicolegal applications.

### Bilateral asymmetry of percutaneous bones of upper and lower limbs

All the long bone lengths for the males showed bilateral asymmetry while for the females, with the exception of radial length, bilateral asymmetry was observed in the remaining limb dimensions (p < 0.05) (Table 3). Low standard of living, nutritional deficiencies, hardships and increased stress in the environment during the developmental stages of life are potential sources of asymmetry [29,30]. It could be inferred that, probably the aforementioned potentially influenced the differential asymmetry observed in the present study, although that was not taken into consideration during the data collection.

Other similar studies carried out among diverse populations have reported contrasting findings regarding asymmetry of humeral length which showed greater values for the right side than the left [7,31–33]. The conflicting findings could be due to differences in physical activities engaged in by participants which was not accounted for in the present study. The bilateral asymmetry seen in both sexes across the entire study population is consistent with a study by Sume [28], who also reported a difference between the right and left tibiae. Interpopulation variations in body parts may be attributed to genetics, geographic location and nutrition [34,35]. Appreciation of variations in anthropometric parameters across populations is therefore relevant in the designing of appropriate products [18]. This would help to produce best fit materials including prostheses and personal protective equipment peculiar to a particular population for maximum benefit.

The majority of the research participants were right-handed (dextral), which fits with the pattern found in the current study. Although this is possible (as about 90% of the general population are right-handed according to De Kovel *et al.* [36]), handedness was not recorded during the data collection, a potential limitation in this study. Handedness has been speculated to have influence on limbs as in the case of dextral individuals, the incessant use of the right limb could have resulted from the increased cortical area within the brain and thus an enhanced neural activation with the right upper limb [37]. In other words, the opposite side control of the cerebral hemisphere of the limbs could perhaps explain this phenomenon. The left cerebral hemisphere is said to have larger surface area than that of the right, thus has greater functional role to exert its effect on the right limb [7]. The left hemisphere being larger and controls motor activities of the right part of the body might explain the superiority of the left cerebral cortex in terms of function resulting in right limb dominance [38]. Therefore, possibly, the right upper limb parameters were shorter because of the effective contraction of muscles compared to the left side and thus, the observed bilateral asymmetry. Interestingly, it has been opined by Vettivel *et al.* [39] that bilateral asymmetry of upper limb bones could be inherited, therefore explaining some of the variation observed in the study.

### Prediction of percutaneous femoral length using upper limb percutaneous bone lengths

Humeral length correlated positively and moderately with its homologous femoral length on both sides for the males (r _right_ = 0.617; r _left_ = 0.639) and females (r _right_ = 0.578; r _left_ = 0.604) (Table 4). Linear regression analysis revealed the humeral length to be the solitary most useful estimator of femoral length on both sides, although the co-efficient of determination of the left side (adjusted R^2^ for males = 0.404; adjusted R^2^ for females = 0.359) was greater than that of the right (adjusted R^2^ for males = 0.376; adjusted R^2^ for females = 0.328). However, for the females, the regression equation that best predicted femoral length utilized left humeral, radial and ulnar lengths (adjusted R^2^ = 0.591).

Overall, the models for the estimation of femoral lengths were more useful regarding the left side compared to that of the right side based on the adjusted R^2^ values. The degree of predictability of the equations improved when other parameters were utilized in the prediction, suggesting that multivariate equations are more useful than single variable predictors. Findings of the study has revealed sex and laterality is useful regarding the use of percutaneous long bones of upper limb for estimation of lengths of percutaneous femoral length. The differences in predictive variables and adjusted R^2^ among the different sexes could be attributed to probable hormonal variations and socioeconomic differences since they may have effects on interstitial (longitudinal) growth of the bones. Again, nutrition and some disease states during childhood period peculiar to the study populations could explain some aspects of the dichotomy. The models derived would serve useful purposes for the orthopaedic surgeon during the reconstruction of the femur using the appropriate upper limb parameters.

### Prediction of percutaneous tibial length using upper limb percutaneous bone lengths

Radial length correlated weakly with its homologous tibial length on the right side (r = 0.270) but moderately on the left side (0.365) among the males. However, among the males, although non-homologous, humeral length correlated moderately better with tibial length on both sides (r _right_ = 0.680; r _left_ = 0.677) (Table 4).

Also, for the females, regarding all the study participants, ulnar length related moderately with its homologous tibial length (r _right_ = 0.658; r _left_ = 0.693). Non-homologous ulnar length correlated moderately with tibial length for the right side (r = 0.649), but strongly for the left side (r = 0.724) (Table 4). For the males, regardless of laterality or homology, the regression equation that best predicted tibial length utilized left humeral and ulnar lengths (adjusted R^2^ = 0472) whereas that of the females, it was left ulnar, radial and humeral lengths (adjusted R^2^ = 0.636). These varied results support the reason why the hypothesis behind the prediction of tibial length from the upper limb percutaneous long bone lengths based on sex is as a result of the existence of interpopulation variations in various body parts due to prevailing environment and genetics. That is why models peculiar to a particular population are recommended owing to the fact that, they are associated with reduced errors.

### Prediction of percutaneous fibular length using upper limb bones

For the males, ulnar length correlated moderately with its homologous fibular length on both sides (r _right_ = 0.519; r _left_ = 0.541). However, among the males, although non-homologous, humeral length correlated strongly with fibular length on the right side (r = 0.716) but moderate on the left side (r = 0.692) (Table 4). For the females, ulnar length correlated moderately with its homologous fibular length on the right side (r = 0.664), but strongly on the left side (r = 0.735). This means that the left parameters showed better associations than that of the right.

Among the males, regression analysis revealed right humeral length and ulnar length to be the most useful estimators of right fibular length (adjusted R^2^ = 0.546), while that of the females were left ulnar length, left humeral length and left radial length (adjusted R^2^ = 0.632). These differential findings further affirm the need for sex specific models to mimimise errors in prediction. The very low standard error of estimates obtained for the models make the regression relations acceptable for safe utilization [40]. Moreover, most of the estimated mean percutaneous lower limb bone lengths using the derived models were not significantly different from that of the actual measured values.

## Conclusion

Males recorded significantly greater percutaneous long bones for both the upper and lower limbs than the female participants. With the exception of radial length among the females, for both sexes bilateral asymmetry was observed in the remaining percutaneous limb dimensions where the upper limb parameters were greater on the left side than the right side, but showed right dominance for the lower limb parameters. Femoral length was best predicted by left humeral length among the males but left ulnar length, left radial length and left humeral length among the females. For tibial length, among the males, it was best estimated by left humeral length and left ulnar length but left ulnar length, left radial length and left humeral length for the females. Meanwhile, for fibular length prediction, among the males, the best estimators were right humeral length and right ulnar length but for the females, they were left ulnar length, left humeral length and left radial length. Although several equations were derived, most of them utilized humeral length in the prediction of lower limb length estimation.

Findings of the present study are useful for the biological profiling of humans with dismembered body parts involved in various disasters such as automobile accidents. The formulae derived would be useful for the design of prostheses. This study has reaffirmed the existence of sexual and bilateral dimorphism in body dimensions.

## Data Availability

Data used to support the study findings would be made available by the corresponding author upon request.

## Acknowledgements

We are much grateful to all our study participants for willingly partaking in this study.

## Author Contributions

Conceptualization: Daniel Kobina Okwan, Chrissie Stansie Abaidoo, Pet-Paul Wepeba, Juliet Robertson, Samuel Kwadwo Peprah Bempah

Data curation: Daniel Kobina Okwan, Priscilla Obeng, Ethel Akua Achiaa Domfeh, Sarah Owusu Afriyie, Thomas Kwaku Asante

Methodology: Daniel Kobina Okwan, Chrissie Stansie Abaidoo, Pet-Paul Wepeba, Juliet Robertson, Samuel Kwadwo Peprah Bempah, Priscilla Obeng, Ethel Akua Achiaa Domfeh

Formal analysis: Daniel Kobina Okwan, Chrissie Stansie Abaidoo, Pet-Paul Wepeba, Juliet Robertson, Samuel Kwadwo Peprah Bempah, Sarah Owusu Afriyie, Thomas Kwaku Asante

Resources: Daniel Kobina Okwan, Chrissie Stansie Abaidoo, Pet-Paul Wepeba, Thomas Kwaku Asante

Validation: Chrissie Stansie Abaidoo, Pet-Paul Wepeba, Daniel Kobina Okwan, Thomas Kwaku Asante

Writing (initial draft/editing/reviewing): Daniel Kobina Okwan, Chrissie Stansie Abaidoo, Pet-Paul Wepeba, Juliet Robertson, Samuel Kwadwo Peprah Bempah, Priscilla Obeng, Ethel Akua Achiaa Domfeh, Sarah Owusu Afriyie, Thomas Kwaku Asante

